# Effects of the COVID-19 Pandemic on Park Use in U.S. Cities

**DOI:** 10.1101/2021.04.23.21256007

**Authors:** Jonathan Jay, Felicia Heykoop, Linda Hwang, Jorrit de Jong, Michelle Kondo

## Abstract

**Introduction:** The COVID-19 pandemic focused attention on city parks as important public resources. However, it is unknown how city park use in 2020 compared to prior years and whether COVID-19 may have exacerbated racial/ethnic inequities in access. Moreover, traditional methods of measuring park use present major drawbacks.

**Methods:** We analyzed monthly mobility data derived from a large panel of smartphone devices, cross-referenced with a database of parks locations sourced from local agencies. We assessed park use trends in 44 of the 50 most populous U.S. cities from January 2018 to November 2020 using interrupted time series regressions. We also compared parks to other city amenities (*e*.*g*., gyms and libraries).

**Results:** Based on a sample of 5,559 city parks, park visits declined by 14.6% (95% CI [9.2, 19.7], p < 0.001) from March through November 2020, compared to prior levels and trends. When we segmented the COVID-19 period by time of widespread closures (March-April) and partial-to-full reopenings (May-November), we estimated a larger reduction during closures (35.7% reduction, 95% CI [33.5, 37.8], p < 0.001) compared to the reduction during reopenings (8.0% reduction, 95% CI [1.9, 13.7], p = 0.001). Reductions for other amenities were more prolonged. In park service areas where a greater proportion of residents were White, reopening was associated with more visits, suggesting that racial privilege influenced access.

**Conclusions:** Smartphone mobility data can address a data availability gap for monitoring park use. Park use only declined modestly in 2020. Opportunities exist to make access more racially equitable.

**Significance statement:** Parks are public resources that promote health. Little is known about how parks have been used during the COVID-19 pandemic, when parks have become particularly important public spaces. This study introduces an approach to monitor park use over time, using location data from smartphones. This approach enabled the authors to evaluate trends in park use during the pandemic, including major gaps in visits to parks according to whether they mostly served White residents or residents of color. This big data approach offers advantages over traditional methods for monitoring park use and can help city officials to identify and address inequities in park access.

## Introduction

Parks are public resources that promote health by providing spaces for exercise and play, access to nature, protection from heat, and opportunities to socialize and build community cohesion. These functions may be particularly valuable in large cities, where higher residential density increases the value of access to outdoor greenspace. During the COVID-19 pandemic, indoor social gatherings and other recreational activities carried serious risk of disease transmission in most communities. Additionally, other amenities for sports and leisure were forced to close. Parks represented comparatively low-risk alternatives. As a result, park use is reported to have boomed in 2020.^1^ On the other hand, we might expect increases in city park use associated with changing exercise and leisure patterns to be offset by drops associated with restrictions on organized sports, the cancellation of concerts and other large gatherings, and closures of schools and playgrounds. This article introduces an application of smartphone data to describe and analyze park use in 2020, compared with previous, non-pandemic years.

In particular, we are interested in disaggregating those data by race: did Black, Latinx, and other residents of color use parks at the same rates as non-Hispanic White residents in 2020? People of color (“POCs”) have experienced a disproportionate burden of COVID-19 infection and mortality,^2^ which may have increased their caregiving demands^3,4^ or influenced their perceptions of safety in public spaces.^5^ Since U.S. communities are characterized by high rates of residential racial segregation, public spaces in majority-POC communities may indeed have carried elevated COVID-19 transmission risk.^6^ Moreover, POCs are disproportionately represented among essential workers and less able to work from home, which may reduce their time available for recreation.

Racial and ethnic disparities in parks access existed prior to COVID-19. POCs have reported barriers to park use including a relative lack of leisure time,^7,8^ lack of social support to use parks for exercise,^8,9^ and lack of programs that suit needs of the community such as afterschool programs or fitness programs outside of traditional workday hours.^9,10^ Although urban-dwelling POCs typically live in areas with higher concentrations of parks within walking distance (*i*.*e*., 0.5 miles, or approximately a 10-minute walk for an able-bodied resident) compared with non-Hispanic white residents,^11–15^ they report walking routes that are unsafe or poorly maintained.^12,13,15,16^

Little empirical research has measured changes in park use during COVID-19. We conducted empirical tests of these hypotheses: (1) Park use in U.S. cities increased in 2020, compared to prior years, and (2) Trends in city park use differed in 2020 by race. Traditional approaches for measuring park use would severely limit our ability to test these hypotheses. Systematic observation of parks^10,17^would rely on the availability of pre-COVID observations and pose difficulties in conducting in-person data collection on a large scale. Population surveys, assuming they could be compared to pre-COVID results, would be limited in their utility by self-report.

Digital methods may offer the most suitable combination of objectivity and scalability. Some prior work has captured park use based on geolocation data from cohorts using wearable trackers^16,18–20^ or from geotagged social media posts.^14^. For this study, we use parks visit data generated from the locations of a sample of smartphone devices equal to approximately 10% of the U.S. population. While these mobility data have been used extensively to examine COVID transmission dynamics,^21–24^ their application to understanding other health behaviors has been limited. Prior work using smartphone mobility data found that U.S. park visits declined substantially during the initial weeks of the COVID-19 pandemic.^23^ Another study found that park use rebounded considerably after that initial drop, but since the authors used data indexed to the initial weeks of 2020, they were unable to assess whether the rebound was atypical compared to ordinary seasonal trends in park use.^25^

Here, we employ monthly panel data from SafeGraph on visits to a large sample of city parks, drawn from 44 large U.S. cities, with observations from January 2018 through November 2020. Since raw SafeGraph data pose challenges for analyzing longitudinal trends in place visits, particularly for parks, we propose methods for matching SafeGraph parks to validated parks data and generating longitudinal comparisons of park use. We use these methods to estimate changes that occurred in 2020, beginning with the initial wave of shutdown measures in March. Beyond testing the hypotheses about increases park use and differences in use according to race, we explore the potential of the data and methods used to monitor the use of local amenities in real time and disaggregate in meaningful ways that can inform city policies.

## Methods

### Data

Our primary data source for this study was monthly park visit panel data provided by SafeGraph, a company that aggregates data on smartphone device locations from users whose settings allow their location data to be collected anonymously by 3^rd^ party applications. The data we used is aggregated by points of interest (“POI”), each of which represents a spatial location (*e*.*g*., a coffee shop at a particular address) where a device could be located when it issues a “ping” to SafeGraph. Although SafeGraph maintains a database of visits to over 4 million POIs, these locations are predominantly commercial locations. In principle, measuring foot traffic at parks is not more challenging than measuring foot traffic at retail stores; however, there may be limited incentives for for-profit data companies like SafeGraph to maintain park visitation data that are as complete and accurate as retail locations data. A 2020 analysis found that the SafeGraph parks data for Philadelphia corresponded to roughly half of the city’s parks acreage and that the polygons SafeGraph used to represent parks boundaries sometimes overlapped only partially with the actual park boundaries.^26^

To address these shortcomings, we cross-referenced the SafeGraph park locations with park locations assembled by the Trust for Public Land (TPL). TPL includes publicly-accessible parks, trails, and open space in its database.^27^ The TPL park database is based on city-, town-and community-provided parks data or, when no data was provided, park data was created based on available resources such as park information from municipal websites, spatial data available from counties and states, and satellite imagery. We considered a SafeGraph park to have matched a TPL park if the SafeGraph park centroid intersected a TPL polygon and the area of the SafeGraph polygon was within 0.5 to 1.5 times the area of the TPL polygon. SafeGraph parks that matched TPL parks (hereafter, the “SafeGraph sample”) were included in the study. This process was designed to omit SafeGraph parks with poorly-drawn polygons or which corresponded to places (*e*.*g*., outdoor museums) that local agencies do not consider to be parks. Also, we included only parks that have appeared in the SafeGraph dataset every month since January 2018, omitting a substantial proportion (approximately 1/3) that were added in mid-2020, and securing a full panel across the study duration. We conducted data processing and analyses in R. Additional technical details and replication code are included in the **Appendix**. We applied this technique using all parks in each dataset within the geographical boundaries of each of the 50 most populous U.S. cities, discarding observations from 5 cities that did not provide geographical boundary data online and 1 city (Colorado Springs, CO) where fewer than 25 SafeGraph parks matched TPL parks. The resulting dataset included park visits from 44 cities, ranging in population from over 8 million (New York City, NY) to approximately 400,000 (Arlington, TX).

We also obtained area-based estimates of the characteristics of residents living within a 10-minute (or 0.5 mile) walk of each park, or within service areas delineated within TPL’s ParkServe database. These estimates were calculated by creating a 10-minute walkable service area using a nationwide walkable road network dataset provided by Esri, and using these service areas to generate overall access statistics for each park, place, and urban area included in the database, and then further disaggregating by race/ethnicity, age, and household income. A 10-minute walking distance is frequently cited as the service area of city parks. We used these data to assess the representativeness of parks included in the SafeGraph sample and calculate sampling weights equal to the inverse of the probability that each park would be included in the SafeGraph sample, given its service area characteristics and the city where it was located.

Our outcomes were the total number of monthly visits to each park in the SafeGraph sample, from January 2018 through November 2020. Following recommendations from SafeGraph, we adjusted these raw visit totals to account for changes in the size of the panel of smartphone devices in each city from month to month. For the cities we analyzed, the total panel size ranged from approximately 4.5 million to approximately 10.2 million over the months we studied. Although SafeGraph recommends removing outlying observations, we manually reviewed the highest-visit observations and believed these counts to be plausible. For example, the farthest outliers were visits to Balboa Park in San Diego during the month that the city hosted the ComicCon convention close to the park. We accounted for the statistical influence of outliers using our regression technique (see below). Finally, over time, recorded visits to POIs tend to increase because SafeGraph’s systems improve at recording these visits, rather than any underlying change in device behaviors; we account for this exogenous effect using model parameters.

To contextualize our park use findings, we obtained SafeGraph data on visits to other public amenities used for exercise (gyms) and social connection (places of worship) and which represent considerable public investments (libraries and transit stations). For these comparator locations, we followed the same data preparation procedures as for park visits, except we assumed that all points of interest were correctly bounded and classified, omitting the cross-referencing step.

### Statistical analysis

Our unit of analysis was monthly observations for each park in the sample. Our main model was an ordinary least squares (OLS) regression with calendar-month fixed effects, to account for seasonality. To remove bias from increased recording of POI visits (as discussed above), we included calendar-year fixed effects. Since the visit counts were distributed approximately log normal, we modeled the natural log of our outcome variable, which also limited the influence of outliers. We applied sampling weights, described above, to each model. We report our results as changes in the rates of park visits. Since intraclass correlation (in our case, observations that might be correlated within cities, *e*.*g*. due to weather patterns or local transmission dynamics) can bias standard errors, we clustered our standard errors by city.

We used an interrupted time series framework to examine how societal changes during COVID-19 influenced park visits, compared to prior trends. The vast majority of U.S. jurisdictions implemented some closure measures in March 2020 and some reopening measures in May 2020.^28^ No U.S. city was COVID-free by the end of November 2020. While the specific timing of closures had only modest effects on population mobility,^29^ changes in daily activity patterns roughly tracked the timing of different policy measures. Conceptually, we hypothesized that COVID-related effects would function as shocks that changed the level of visits without changing the month-to-month slope, which otherwise would be governed by seasonal trends.

We tested the impact of three sets of exposures: (1) all COVID-related effects, as measured by a binary indicator that switched on from March 2020 through November 2020; (2) separate closure-related effects (March-April 2020) and reopening effects (May-November 2020), using binary indicators; and (3) the moderating effect of racial privilege on parks access, as measured by the interaction of the closure and reopening indicators with a continuous variable representing the proportion of residents in the service area population of each park who were White (Hispanic or non-Hispanic). We did not observe closures or reopenings of specific parks in our sample, but rather assigned indicators for closure and reopening time periods based on the national trends in public policy and population mobility that prior work has identified. We used likelihood ratio tests to assess the change in fit as we added terms to each model.

We recalculated the first model (*i*.*e*., COVID-related effects) separately for each city to characterize city-level variation. We also recalculated the second model (*i*.*e*., closure vs. reopening effects without moderation) using comparator location visits as the outcome variable. All input parameters were the same except for model weights, which were used to give equal weight to each amenity category (*i*.*e*., gyms, places of worship, libraries, transit stations), since these categories varied widely in the number of points of interest available in the SafeGraph data.

## Results

Of the 19,115 parks in the Trust for Public Land database that fell within city boundaries, 5,630 (29%) matched a park in the SafeGraph data. **Table 1** displays the sample characteristics. Matched parks displayed higher acreage and served a larger population that comprised a larger proportion of White residents compared to all parks within city boundaries. The proportion of older residents and low-income households in the service area for matched parks were similar to those of other city parks.

**Table 1:** Characteristics of parks sample

|  | All city parks | Parks matched in<br>smartphone data |
| --- | --- | --- |
| *N* | 19115 | 5559 |
| Acres | 19.5 (112) | 21.4 (93.2) |
| Total population within 10-min walk | 6223 (9157) | 6210 (9207) |
| Low income household %<br>within 10-min walk | 39.6 (20.8) | 39.9 (19.3) |
| White population % within 10-min walk | 50.3 (28.2) | 54.4 (26.2) |
| Senior population % within<br>10-min walk | 13.5 (6.5) | 14.1 (6.0) |

Figure 1 depicts trends in park visits after accounting for sampling probability, changes in the size of the SafeGraph panel, and between-year differences in visit recording. When we examined the effects of COVID-19 from March through November, we found a 14.6% reduction in park visits (95% CI [9.2, 19.7], p < 0.001) compared to prior trends. **Table 2**. Most cities displayed point-estimated reductions between 10% and 30%, as displayed in **Supplemental Figure 1**.

**Figure 1:**
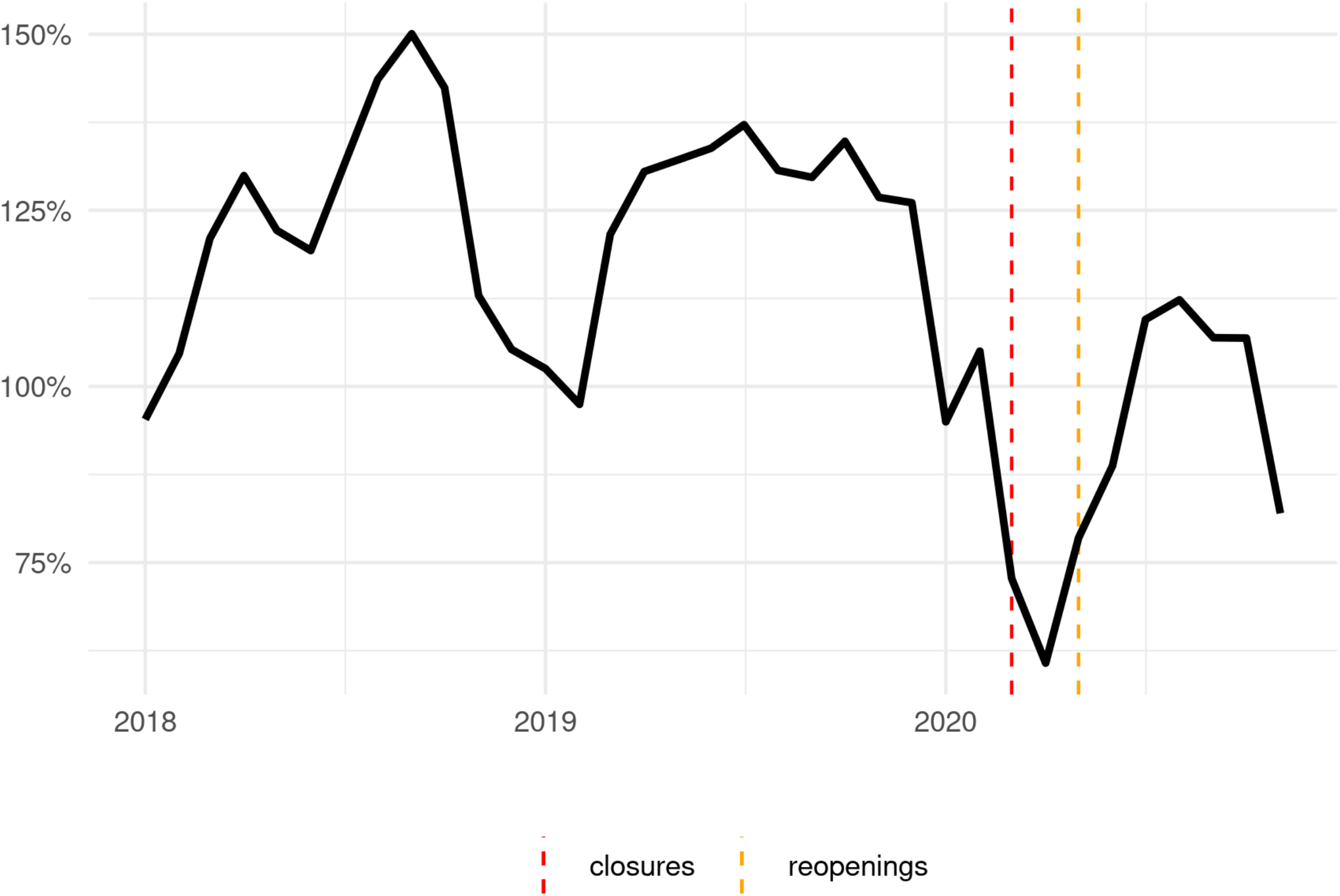
Monthly park visits, scaled by visits in January-February of the same year.

**Table 2:**
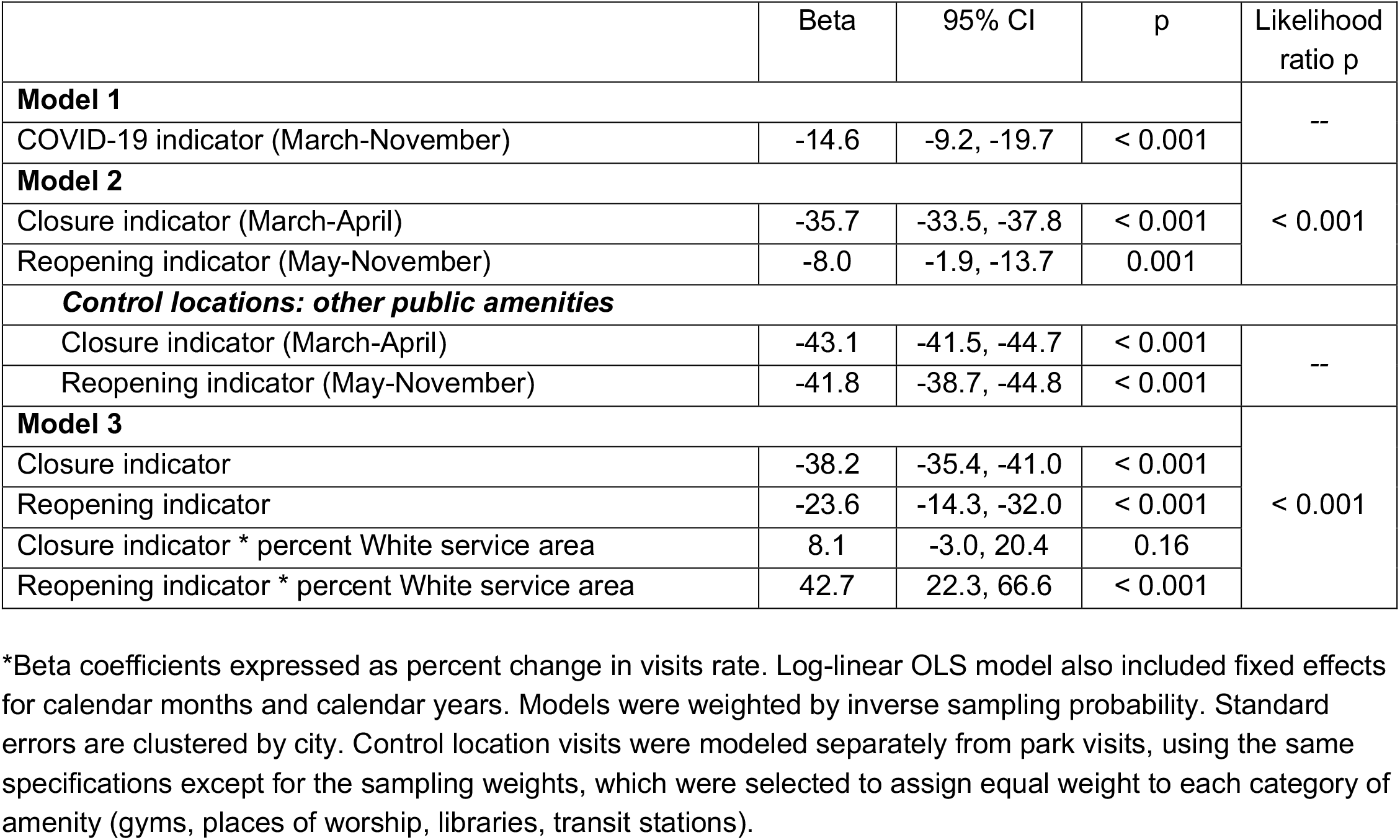
Estimated effects of COVID-19 on park visits from regression models

|  | Beta | 95% CI | p | Likelihood ratio p |
| --- | --- | --- | --- | --- |
| **Model 1** |  |  |  | -- |
| COVID-19 indicator (March-November) | -14.6 | -9.2, -19.7 | < 0.001 |  |
| **Model 2** |  |  |  | < 0.001 |
| Closure indicator (March-April) | -35.7 | -33.5, -37.8 | < 0.001 |  |
| Reopening indicator (May-November) | -8.0 | -1.9, -13.7 | 0.001 |  |
| ***Control locations: other public amenities*** |  |  |  |  |
| Closure indicator (March-April) | -43.1 | -41.5, -44.7 | < 0.001 | -- |
| Reopening indicator (May-November) | -41.8 | -38.7, -44.8 | < 0.001 |  |
| **Model 3** |  |  |  | < 0.001 |
| Closure indicator | -38.2 | -35.4, -41.0 | < 0.001 |  |
| Reopening indicator | -23.6 | -14.3, -32.0 | < 0.001 |  |
| Closure indicator * percent White service area | 8.1 | -3.0, 20.4 | 0.16 |  |
| Reopening indicator * percent White service area | 42.7 | 22.3, 66.6 | < 0.001 |  |

\*Beta coefficients expressed as percent change in visits rate. Log-linear OLS model also included fixed effects for calendar months and calendar years. Models were weighted by inverse sampling probability. Standard errors are clustered by city. Control location visits were modeled separately from park visits, using the same specifications except for the sampling weights, which were selected to assign equal weight to each category of amenity (gyms, places of worship, libraries, transit stations).

When we segmented the COVID-related effects in the full sample, we found a 35.7% reduction associated with closures (95% CI [33.5, 37.8], p < 0.001) and an 8.0% reduction associated with reopenings (95% CI [1.9, 13.7], p = 0.001). This modification improved the model fit (p < 0.001). **Figure 2** compares trends in park visits to other public amenities. In our models, visits to comparator locations declined by a slightly larger margin during closures than park visits did (43.1% reduction, 95% CI [41.5, 44.7], p < 0.001), but unlike park visits, they displayed virtually no increase in visits during reopenings compared to prior years (41.8% reduction, 95% CI [38.7, 44.8], p < 0.001). See **Table 2**.

**Figure 2:**
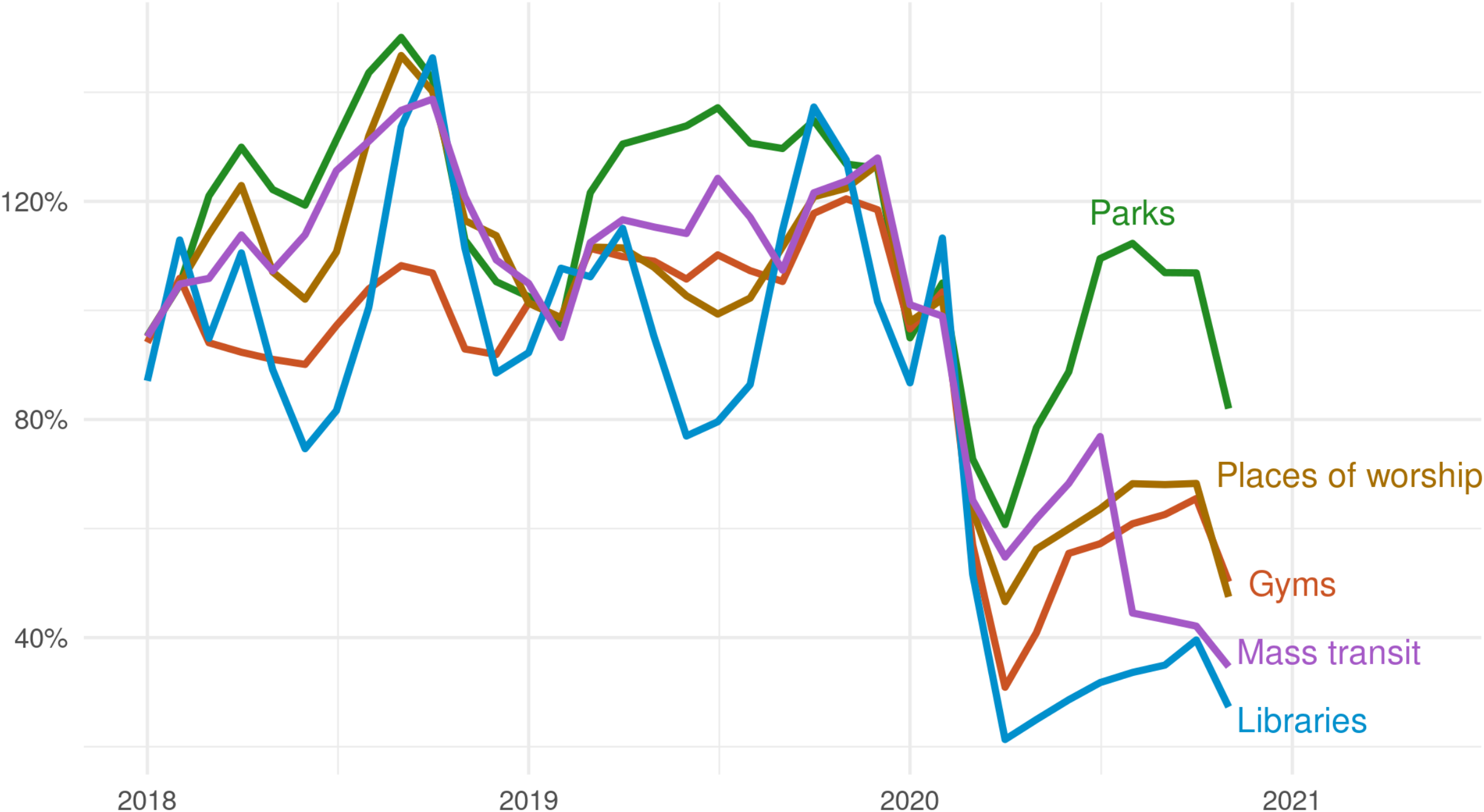
Monthly visits, scaled by visits in January-February of the same year, for parks and other public amenities.

Figure 3. compares trends in visits at parks serving majority-POC and majority-White populations. In the model that tested the moderating effect of White population served, we found that White population did not moderate closure effects but moderated reopening effects considerably. A 1-unit change in the moderator (*i*.*e*., from a 0% White to a 100% White service area) was associated with 42.7% greater park visits (95% CI [22.3, 66.6], p < 0.001) post-reopening. See **Table 2**. Consequently, at the highest levels of White population served, the model estimated that visits in the reopening period were greater than in the same time period in prior years. **Supplemental figure 2** displays these marginal effects. The moderated exposure model improved fit compared to the previous model (p < 0.001).

**Figure 3:**
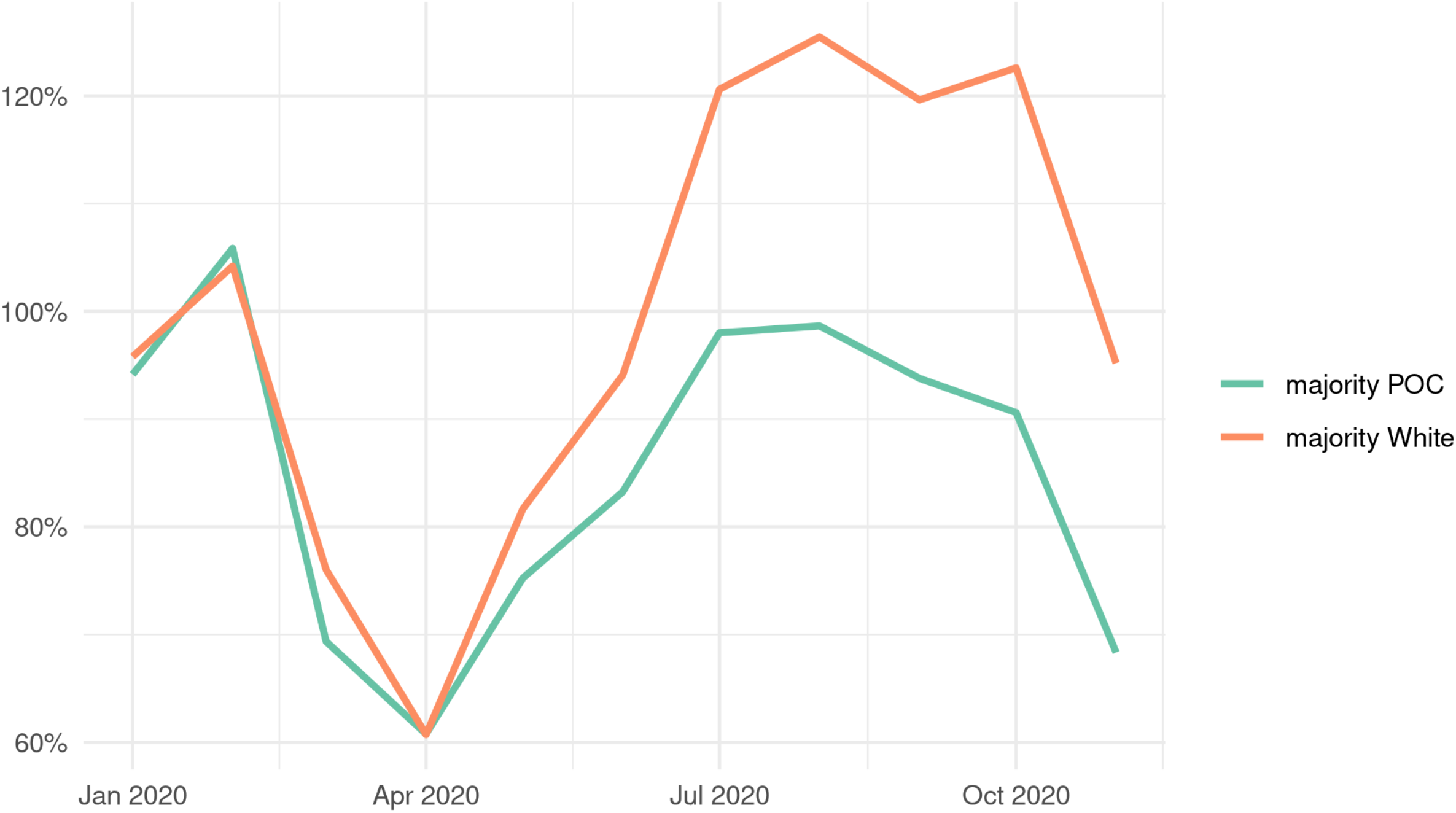
Monthly park visits, scaled by visits in January-February of the same year, by service area racial composition.

## Discussion

Using data from a large sample of city parks in major U.S. cities, we found that total park use during COVID-19 was lower than the levels observed in 2018 and 2019. It appears that this reduction was larger during the months in which most public spaces were closed; after jurisdictions began reopening, the reduction in park visits was statistically significant but fairly modest. This pattern contrasted with visits to other public amenities, which did not rebound substantially after jurisdictions began reopening. However, for parks, the effect of reopening varied substantially according to the racial composition of the population living within walking distance. Where the proportion of White residents was higher, reopening entailed a larger rebound in park visits, including an overall *increase* in visits, compared to 2018 and 2019, in predominantly White service areas.

These findings highlight, first, the valuable function that city parks occupied during the COVID-19 pandemic. During a period when other public amenities were underutilized and population mobility declined substantially, city park use was reasonably steady following the March-April drops. Despite parks’ apparent value to city residents, the economic fallout caused by the pandemic poses fundamental challenges to city parks departments: in 2020, 38% of U.S. mayors reported that they expect “dramatic financial cuts” to parks and recreation budgets in their cities because the fiscal crisis forces cities to cut spending.^30^

Second, our analysis suggests that structural racial inequities shaped city residents’ ability to visit parks during COVID-19. During a period when parks represented comparatively safe venues for exercise, recreation, and outdoor gatherings, parks that served predominantly White populations were well-utilized compared to their baseline levels. This finding is consistent with media reports on high rates of park use during COVID-19, potentially underscoring how media narratives center White experiences.^1,31^

By contrast, we found a relatively small rebound in park visits, after initial closures, in parks serving a larger proportion of residents of color. Our study design does not enable us to evaluate the possible causes of these patterns. Possible causes include disparities in COVID-19 infection and mortality, disparities in perceived risk to self or others, disparities in economic distress, or disparities in access to parks. Moreover, 2020 was marked with highly visible instances of interpersonal and structural racism, including the police killings of Breonna Taylor, George Floyd, and other unarmed Black people, which might have influenced how residents of color perceived their safety in public spaces, including parks. These questions are important topics for future research and for local policymakers.

Finally, we demonstrated the utility of smartphone mobility data for tracking time trends in city park use. This application of a novel data source addresses a major gap for city officials, researchers, and other stakeholders attempting to monitor park use over time. This approach enabled us to assess time trends across dozens of cities, with high temporal resolution, using a sample of parks that was hundreds of times larger than traditional observational methods have used. We have made the longitudinal trends from a subset of cities in this sample available at [LINK]. More granularized methods, which others have advanced,^32^ could potentially allow local actors to estimate actual park visits in close to real time, enhancing their ability to identify barriers to access and evaluate program effectiveness. Moreover, SafeGraph has provided these data to researchers and public officials at no cost; even at standard pricing, the data would likely remain highly cost-effective compared to in-person data collection.

### Limitations

SafeGraph park use data have not been validated against traditional data sources for park use, such as systematic observation. However, prior work has shown that SafeGraph park use data track closely with Google Community Mobility Reports for park use, which are derived from smartphone locations using different data collection methods.^23^ SafeGraph device visits are difficult to interpret in terms of real-world visitors, particularly after adjusting for changes in the size of the SafeGraph panel. Therefore, our analyses here focus solely on time trends. In other words, we do not attempt to compare the *absolute* levels of park use by population served, but only how the time trends differed according to population served.

Moreover, we were not able to assess the manner in which city parks were used, or the characteristics of the persons using them. Because we could not observe directly the racial/ethnic identity of park users, we assigned these characteristics at the park level based on the demographics of the population living within a 0.5 mile walk. Park users might travel substantially farther than this distance,^33^ but prior work also suggests that park users are likely to seek out venues in neighborhoods where the demographic composition is comparable to their own.^34^ Therefore, we considered nearby demographics to be a suitable proxy for park user characteristics. Still, as noted above, we do not draw conclusions about the precise cause of our findings relating to the use of majority-White serving parks, which could conceivably include people of color choosing more often to visit parks in majority-White areas (though we have not found evidence to support this proposition). Future research could examine this question. Moreover, since our racial composition indicators did not include mutually exclusive Hispanic/non-Hispanic categories, we were unable to disaggregate how effects varied depending on White Hispanic and White non-Hispanic populations served.

While we have treated COVID-19-related changes as the exposures in this study, we are unable to assess which causal mechanisms related to COVID-19 may explain the patterns we have observed. We refer to “closure” and “reopening” periods based on the general trends in public policy and population mobility that occurred during certain monthly of 2020, rather than policies that closed or reopened specific parks in our sample, which we did not observe. Future work could extend our methodology to distinguish among the effects on park use from differences in state and local policy, parks characteristics, COVID transmission patterns, and other possible influences.

### Conclusions

The COVID-19 pandemic sharpened focus on city parks as important public resources. However, cities lack adequate tools to monitor and evaluate the use of parks. Our analysis shows that, despite some limitations, smartphone data can be used to learn how park use varies over time and between geographies, and how changes in park visits compare to visits to other amenities. Finally, the data can help identify racial inequities in park access.

As cities are grappling with fiscal challenges in the wake of the pandemic and the subsequent economic fall-out, they need to make difficult decisions regarding allocation of resources. The approach to studying park use developed in this study can contribute to the discussions and decisions about investments in parks. Efforts to enhance community participation,^35,36^ secure sustainable funding streams,^37,38^ and monitor equity^39^ can assist in this endeavor.

In 2020, many cities also began to reckon more deeply with longstanding racial injustice. This study has also demonstrated how cell phone data can be used to identify racial disparities in park use and park access. Additional research is needed to identify the sources and consequences of racial inequities in park visits. While we propose methods that may enable cities to quantify changes in park utilization over time, participatory and qualitative methods are necessary to investigate the causes and chart the path forward. In combination, these strategies could make city parks programming and other parks investments more effective, efficient, and equitable.

## Data Availability

Trust for Public Land data are publicly available at https://www.tpl.org/parkserve/downloads. Raw SafeGraph data cannot be shared due to data use agreement, but replication code will be posted prior to publication.

## Acknowledgements

The authors thank SafeGraph for making data available for this study. They thank Elijah de la Campa, Lara Miller, and Pete Aniello for helpful comments and discussions related to the study and study manuscript. Jordan Boulay, Andrea Kuriyama, Rachel Martin, Manish Patel, and Kristal Xie contributed to data collection.

