## Supplemental Appendix for "Effects of the COVID-19 Pandemic on Park Use in U.S. Cities"

**Supplemental figure 1:** COVID-related changes in park use, by city

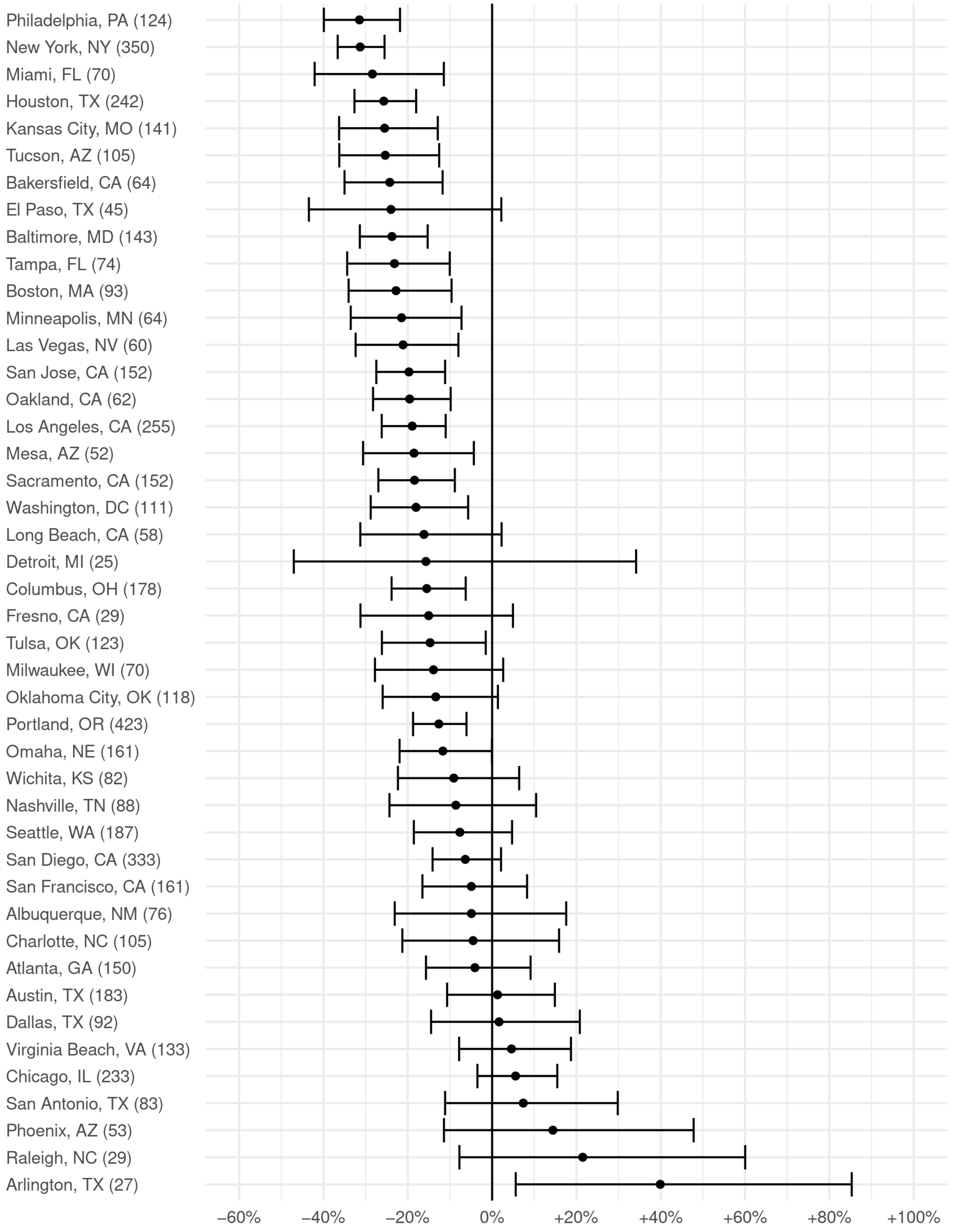


Depicted: point estimates and 95% confidence intervals. The number of parks included in sample for each city is displayed in parentheses. Plot depicts output of Model 1 calculated separately for each city. Model includes log-transformed visits with year and month fixed effects and is weighted by sampling probability. All specifications are identical to Model 1 (see **Table 2**) except that standard errors are not clustered by city.

**Supplemental figure 2**: Estimated marginal effects of proportion White residents and reopenings indicator on park visits

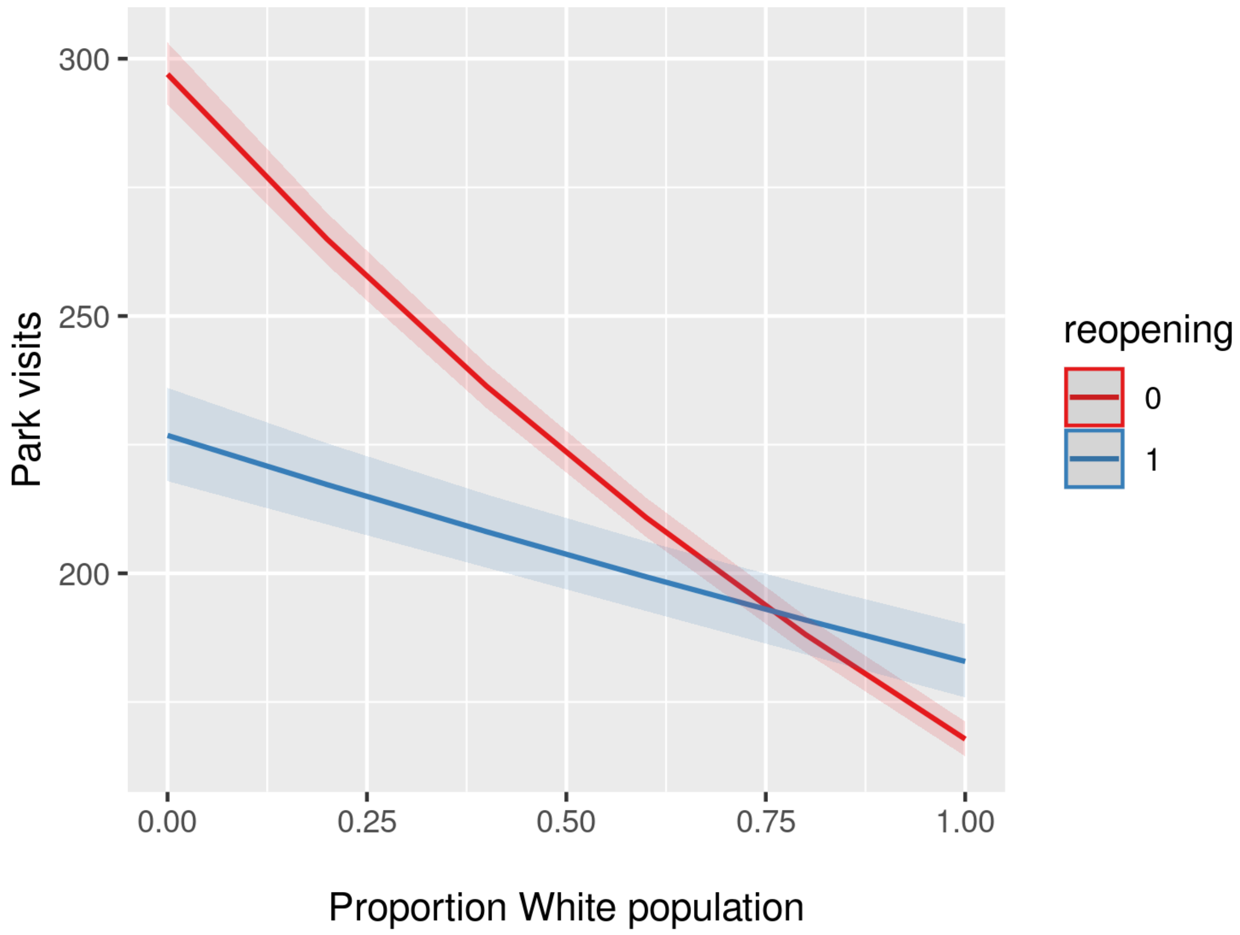


Parks visits measure is raw device visit counts appearing in the SafeGraph dataset. Marginal effects are derived from Model 3 (see **Table 2**).
